# Human blood-brain tumor barrier on a chip to investigate personalized treatment for glioblastoma patients

**DOI:** 10.64898/2025.12.01.25341181

**Authors:** Minsu Ryoo, Gaeun Lee, Jinwoo Jung, Sujin Cho, Sharon Jeeho Ham, Nayeong Kang, Hyeongjin Ahn, Yu Jin Kim, JeongMin Sim, Jeongman Park, Juwon Kim, Sohyun Hwang, Youn-Jung Kang, Jaejoon Lim, Jungho Ahn, Song Ih Ahn

## Abstract

The inherent characteristics of glioblastoma (GBM), including tumoral heterogeneity and invasive capacity, combined with the presence of the blood-brain tumor barrier (BBTB), present challenges in developing effective treatment for GBM. Especially, the margins of GBM, where GBM cells infiltrate normal brain tissue, exhibit high resistance to therapies. Despite the difficulties in controlling tumor progression from this region, the GBM margin remains a critical area to be studied. Here we report a microengineered model that mimics the BBTB within the GBM margin, incorporating a 3D network of normal astrocytes and GBM cells isolated from patients newly diagnosed with GBM. The interaction between GBM cells and stromal cells results in increased vascular permeability, reactive gliosis, and alterations in astrocyte behavior to foster tumor invasiveness and progression. We compare patient-specific tumor responses to conventional chemotherapy in our BBTB on a chip model with clinical outcomes, demonstrating the capability of the model to predict personalized drug responses. Our BBTB model may serve as a personalized tool to examine the interactions between tumors and normal brain tissue, ultimately facilitating the screening of personalized medicine for GBM treatment.

## Introduction

Glioblastoma (GBM) is the most prevalent and aggressive type of brain cancer, with a median survival of less than 15 months despite multimodal therapy.^[1]^ The poor prognosis of GBM is primarily due to its intrinsic heterogeneity across patients and diffusively infiltrative grown pattern, which hinders complete surgical resection and facilitates recurrence.^[2,3]^ A particularly challenging region is the GBM margin, where invasive tumor cells infiltrate normal brain parenchyma.^[4,5]^ In this region, GBM cells interact dynamically with resident glial cells^[6]^ and are shielded by the blood-brain tumor barrier (BBTB), a pathological extension of the blood–brain barrier (BBB), that retains relatively intact vascular barrier.^[7]^ These features not only restrict drug delivery but also contribute to therapy resistance and tumor progression. Therefore, understanding the cellular and barrier dynamics at the tumor margin is critical for improving therapeutic outcomes in GBM.

The heterogeneity of GBM, both within tumors and across patients, contributes significantly to the inconsistent therapeutic responses, even among patients with favorable prognostic markers such as methylated O(6)-Methylguanine-DNA methyltransferase (MGMT).^[8]^ Although patient’s genomic profiling provides important diagnostic and prognostic insights, it does not fully capture the functional differences in tumor biology, particularly in relation to drug permeability and therapeutic response. These limitations highlight the need for patient-specific models that can recapitulate the microenvironment and vascular barrier characteristics of the GBM margin, and rapidly evaluate individual therapeutic responses.^[9]^

Organ-on-a-chip technology has emerged as a promising platform for personalized medicine screening, by combining patient-derived cells with physiologically relevant microenvironments.^[10]^ This technology has adopted diverse strategies to replicate the GBM features, including the integration of GBM spheroids,^[11,12]^ the simulation of hypoxic conditions,^[13–15]^ or the use of patient-derived cells.^[14,16,17]^ However, few models have successfully recapitulated the integrated features of the BBTB within a patient-specific GBM margin.^[18]^

In this study, we present a microphysiological system that recapitulates the three-dimensional (3D) microenvironment of the BBTB within the GBM margin (**Figure 1A** and **Supplementary Figure S1**). Our model is engineered to reconstruct the brain vascular barrier and perivascular environment of GBM within a double-layered microfluidic device.^[19]^ GBM cells were isolated from three GBM patients with MGMT promoter methylation, a marker predictive of response to chemotherapy with alkylating agents like Temozolomide (TMZ).^[20–23]^ Despite sharing this biomarker, the patient-derived models exhibited distinct vascular phenotypes and drug responses, reflecting inter-patient heterogeneity. Our model enabled functional evaluation of BBTB integrity, drug permeability and treatment efficacy in a patient-specific manner, offering a translational tool to predict prognosis and optimize therapeutic strategies for GBM.

**Figure 1.**
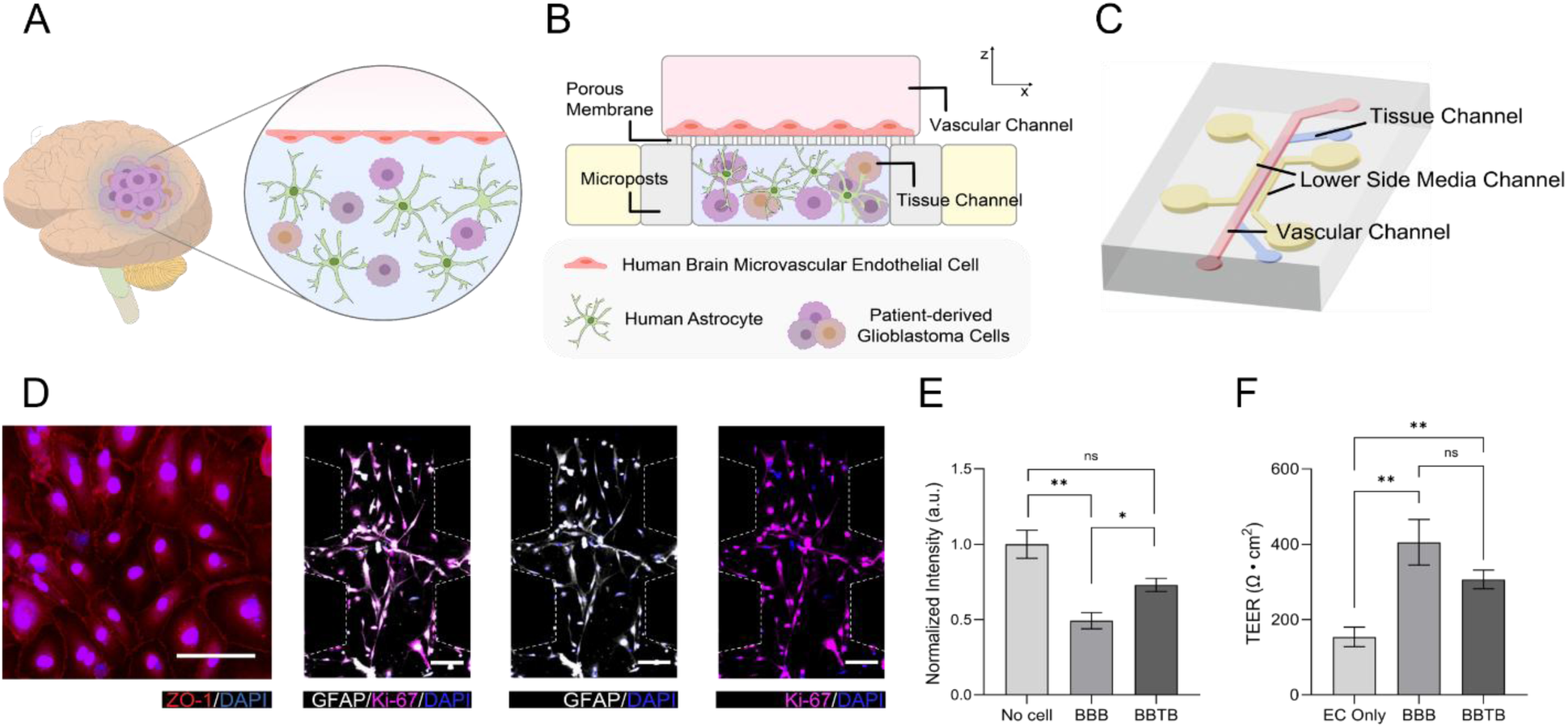
Microengineered human blood–brain-tumor barrier (BBTB) model. **(A)** Schematic representation of the BBTB within the GBM margin. **(B)** Cross-sectional view of our BBTB model consisting of the human brain endothelial barrier in vascular channel (upper channel) and a 3D network of HAs and patient-derived GBM cells in tissue channel (lower channel). The two microfluidic layers are separated by a porous membrane, and the tissue channel and lower side media channel are separated by an array of microposts. **(C)** The microfluidic chip with the vascular channel (red), the tissue channel (blue), and the lower side media channels (yellow). **(D)** HBMEC monolayer formed in the vascular channel (ZO-1, red; DAPI, blue) (scale bar = xx µm). **(E)** Tissue layer showing co-cultured GBM and astrocytes (GFAP, white; Ki-67, magenta; DAPI, blue) (scale bars = xx µm). **(F)** Normalized intensity of tracer movement across the barrier in different experimental conditions: No cells, BBB, and BBTB (n=3 for each condition, *p<0.05, **p<0.01 and **p<0.001 by student’s t-test). **(G)** TEER measurements for endothelial cells only (EC Only), BBB, and BBTB conditions, reflecting the integrity of the barrier (n=4 for EC only, n=3 for BBB, and n=10 for BBTB, *p<0.05, **p<0.01 and ***p<0.001 by student’s t-test).

## Materials and Methods

### Fabrication of the microfluidic device

The microengineered device was fabricated using polydimethylsiloxane (PDMS; Sylgard 184, Dow Corning, Midland, MI, USA) via soft lithography. The PDMS elastomer base was mixed with a curing agent at a 10:1 (w/w) ratio. After degassing to remove air bubbles, the mixture was poured over silicon wafers patterned with SU-8 photoresist (Microfit, Hanam-si, Gyeonggi-do, Republic of Korea) and baked at over 80 °C for 1 hour to allow curing. Subsequently, the solution was poured onto the lower-layer wafer, and spin-coating was applied to achieve a lower channel height of 250 μm. This lower layer was then cured on a hot plate at 130 °C for 18 minutes. Prior to assembly, both the upper and lower layers were cut, and media reservoir holes in the upper layer were punched using a 4 mm biopsy punch (KAI Medical, Seki, Gifu, Japan), while other inlet and outlet ports were created using a 1 mm punch. Both sides of the polycarbonate membrane with 8 μm pores (Sterlitech Corp, Kent, WA, USA) were treated using a vacuum plasma system (Femto Science, Hwaseong-si, Gyeonggi-do, Republic of Korea) for 1 minute. The plasma-treated membranes were then cut into 10 mm × 3 mm pieces and immersed in a 5% solution of 3-aminopropyltriethoxysilane (APTES; Sigma-Aldrich, St. Louis, MO, USA) at 80 °C for 20 minutes. Finally, the membrane was sandwiched between the upper and lower PDMS layers and bonded using the same vacuum plasma system. The assembled device and microchannels were sterilized with 70% ethanol, followed by incubation in a dry oven at 80 °C for over 2.5 days to restore the hydrophobicity of the PDMS surface.

### Glioblastoma cell isolation

The patient tissues were collected from CHA Bundang Medical Center, Republic of Korea, under Institutional Review Board approval (No. CHAMC2021-01-024-001); all patients provided informed consent. Tissues were surgically obtained and stored at −80 °C. The patient sample was diagnosed by a neuropathologist and also underwent next-generation sequencing using the Oncomine™ Comprehensive Assay.

Primary GBM cells were acquired from histological grades 4 GBM tissue samples. Tissues were mechanically sectioned into small pieces and enzymatically dissociated using collagenase D (Roche Diagnostics, Basel, Switzerland) and DNase l (Roche Diagnostics) at 37 °C for 30 min.

The cells were washed with phosphate-buffered saline (PBS; Thermo Fisher Scientific, Waltham, MA, USA) and enumerated using a hemocytometer (Thermo Fisher Scientific). Then, cells, at a density of 1E6 cells/mL, were cultivated in Dulbecco’s modified eagle medium (DMEM) supplemented with 12.5% FBS, 1% (v/v) penicillin/streptomycin, 50 ng/mL epidermal growth factor (EGF; Thermo Fisher Scientific), and 50 ng/µL fibroblast growth factor (FGF; PeproTech, Cranbury, NJ, USA) at 37 °C and in a 5% CO2 atmosphere. All media and reagents were lipopolysaccharide (LPS)-free. FBS was heat-inactivated and contained < 5 EU/mL of endotoxin.

### Cell culture

Human Brain Microvascular Endothelial Cells (HBMEC; Innoprot, Bizkaia, Spain) at passages 3-5 were cultured in endothelial cell medium (Sciencell, San Diego, CA, USA) on flasks coated with 50 μg/mL fibronectin (Sigma-Aldrich). Human Astrocytes (HA; Sciencell) at passages 3-5 were cultured on flasks coated with 1 mg/mL poly-l-lysine (PLL; Sigma-Aldrich) and maintained in astrocyte medium (Sciencell). GBM cells from patients at passage 3-5 were grown in Dulbecco’s Modified Eagle Medium F12 (DMEM/F12, Gibco, NY, USA) with the addition of 50 µg/mL Recombinant Human Epidermal Growth Factor (EGF, Peprotech, Cedarbrook Dr, USA) and 50 µg/mL Recombinant Human Fibroblast Growth Factor-10 (FGF-10 Peprotech) on flasks (**Supplementary Figure S2**).

### Construction of the BBB-GBM chip system

HAs and GBM cells were labeled with CellTracker (CellTracker Red CMTPX Dye, Thermo Scientific; CellTracker Green CMFDA Dye, Thermo Scientific) and then incubated at 37 °C for 30 minutes before seeding into the chips. HAs and GBM cells, with a density of 2E6 cells/mL, were enclosed in a precursor solution of growth factor reduced Matrigel (Matrigel; Corning, NY, USA) at a concentration of 5 mg/mL and loaded in the lower center channel. The cell loaded Matrigel was incubated for 1 hour at 37 °C to allow gelation. After 1 hour, mixed cell culture medium (1:1 of astrocyte medium: endothelial cell medium), was introduced into the lower side channel to provide nourishment to the embedded HAs and GBM cells, preventing Matrigel shrinkage. The chip was subsequently incubated at 37 °C for 24 hours to promote the proliferation of HAs and GBM cells prior to the introduction of HBMECs. Before seeding HBMECs onto the membrane in the upper channel, the channel was coated with 50 μg/mL fibronectin and incubated for 30 minutes at 37°C to enhance the attachment of cells onto the surface of channels. Afterward, HBMECs were seeded into the upper channel at a density of 7E7 cells/mL. After 1 hour, the mixed cell culture medium was replenished in the upper channel to remove unattached cells and supply nutrients to HBMECs. The chips were incubated at 37 ℃ with 5% CO_2_ for 3 days, with the cell medium being refreshed twice a day.

### Immunocytochemistry

Samples were fixed for 15 minutes at room temperature using 2% paraformaldehyde (PFA; Bio-solution, Suwon-si, Gyeonggi-do, Republic of Korea). To prevent contraction of HBMECs, HAs, and GBM cells, the PFA solution was warmed up in a water bath before use. Following the permeabilization of the samples using 0.1% Triton X-100 (Sigma-Aldrich) solution for 15 minutes at room temperature, the samples were blocked using 2% bovine serum albumin (BSA; Sigma-Aldrich) solution for one hour at room temperature.

The lower channels of the samples were treated with primary antibody (rabbit anti-Ki-67 (1:100; Sigma-Aldrich)) and incubated at room temperature for 3 hours, washed three times with 1% BSA and PBS. Subsequently, they were incubated with fluorescence-conjugated secondary goat anti-rabbit AlexaFlour 555 (1:200; Invitrogen) for one hour at room temperature. Afterwards, mouse anti-ZO-1 (AlexFluor 594 conjugated; 1:100; Invitrogen) and mouse anti-GFAP (AlexFluor 488 conjugated; 1:100; eBioscience) were introduced into the upper and lower channels, respectively and incubated for 1 hour at room temperature. Samples were washed three times with 1% BSA and PBS. Nuclei were stained with 1 ug/mL 4′,6-diamidino-2-phenylindole (DAPI; Sigma-Aldrich) in DI water for 5 minutes at room temperature and then washed with DI water and PBS. The stained samples underwent analysis using a confocal microscope (LSM 980, Carl Zeiss, Oberkochen, Germany) at the KAIST Analysis Center For Research Advancement (KARA). The analysis of the images was performed using ZEN 3.1 blue edition software (Carl Zeiss).

### Permeability measurements

After establishing the experimental models, all samples were replenished DPBS (Dulbecco’s Phosphate-Buffered Saline; Welgene, South Korea) before permeability assay. 1 mg/mL of 10 kD FITC-dextran (Sigma-Aldrich) in DPBS was administered into the upper channel. After 10 seconds, 20 μL of solution was sampled from the two reservoirs linked to the lower side channels. The fluorescence intensities of the aspirated samples were measured with a 96-well Microplate reader (Cyto-FLEX-Analyzer). The intensities were normalized against the average of the control group.

### Transendothelial electrical resistance (TEER) measurement

TEER was measured across the endothelial monolayer in each device using a custom-built electrode system. The setup included Ag/AgCl wires (300 μm diameter, 30 mm length) integrated into a modified connector, and linked to an EVOM2 volt-ohmmeter (World Precision Instruments, Sarasota, FL, USA), which applies a 10 μA alternating current at a frequency of 12.5 Hz. To minimize background measurement interference, the electrode wires were encased in Tygon tubing (0.8 mm inner diameter × 2.38 mm outer diameter; Duksan General Science, Republic of Korea) and PE-60 tubing (0.68 mm ID × 1.18 mm OD; Musashi Engineering, Japan), both filled with culture medium. Each pair of Ag/AgCl electrodes was inserted into the inlet and outlet of the upper and lower channels, respectively. Resistance values were recorded after 1 minute stabilization period. For each chip, ten readings were obtained and averaged. Final TEER values were derived by subtracting the resistance measured from blank models and multiplying the result by the endothelial-covered surface area within the lower channel (0.015 cm²).

### Real-time qRT-PCR

HA, GBM cells, and ECs each at a density of 2E5 cells/mL, were seeded into a 6-well plate and a transwell insert (Corning), respectively, and incubated at 37°C for 24 h. Subsequently, the inserts were transferred to the upper compartment. After 48 hours, the culture medium and transwell insert were discarded, and cells were harvested at confluence using 0.25% Trypsin-EDTA (GenDEPOT, TX, USA). Total RNA was extracted using Labozol reagent (CMRZ001, Cosmogenetech, Seoul, South Korea) following the manufacturer’s instructions. Complementary DNA (cDNA) synthesis was performed by converting 1 μg of total RNA using the PrimeScript™ IV 1st strand cDNA Synthesis Mix (6215A, TAKARA, Kusatsu, Japan). RT-qPCR was conducted on cDNA samples using SYBR Green (RT500M, Enzynomics, Daejeon, South Korea) to evaluate the expression levels of target genes. The gene expression data obtained from the experiments were normalized using the housekeeping gene (beta actin; *ACTB*). Primer sequences utilized in the experiments are detailed in **Supplementary Table S1.**

For EC gene expression analysis in patient-specific setups, ECs were seeded in 6-well plates and HAs with GBM cells seeded in transwell insert (Corning) at a total amount of 1.5E5 cells per compartment at 37°C for 24 h. Subsequently, the inserts were transferred to the upper compartment. After 48 hours, the culture medium and transwell insert were discarded, and ECs were lysed in 500 *μL* of Trizol regent (Invitrogen, USA) for subsequent RNA extraction.

Complementary DNA (cDNA) synthesis was performed by converting 1 μg of total RNA using iScript^TM^ cDNA Synthesis kit (Bio-Rad, USA). RT-qPCR was conducted on cDNA samples using SYBR Green (SYBR Supermix; Bio-RAD, Hercules, USA) to evaluate the expression levels of target genes. The gene expression data obtained from the experiments were normalized using the housekeeping gene (*GAPDH*). Primer sequences utilized in the experiments are detailed in **Supplementary Table S2.**

### TMZ treatment and evaluation efficacy

Temozolomide (TMZ; Sigma-Aldrich) reconstituted in DMSO was added to culture media in concentration of 50 uM and 500 uM respectively. To investigate the chemotherapeutic specific response of cancer cells in GBM patients, HA, GBM and EC cells were treated with or without 50 uM and 500 uM TMZ for 72h in a mixture of astrocyte media (AM) and GBM media in a 1:1 ratio. The mixture medium was filled into the upper and lower side channels of the device.

Following TMZ treatment, to measure the cell viability of GBM cells per patient in response to TMZ treatment, HA and GBM cells in the lower channel of the device were stained with 0.5 μg/mL propidium iodide solution (PI; Biolegend, San Diego, CA, USA) in DPBS at room temperature in the dark for 1 h. Subsequently, to prevent nonspecific staining and remove excess PI dye, excess PI staining solution was carefully aspirated, and the cells were washed again with DPBS. The stained cells were imaged using a fluorescence microscope for PI detection in the Texas Red channel. The stained cell area was quantified using ImageJ, and the resulting cell area values were normalized against the average of the control group.

### Statistics analysis

All statistical analyses were performed using GraphPad Prism 10 (GraphPad Software, La Jolla, CA, USA). Comparison groups were analyzed with Student’s t-test. Statistical significance was determined based on the P-values: P<0.05(*), P<0.01(**), P<0.001(***) and P<0.0001(****).

## Results

### Reconstruction of the blood-brain tumor barrier in glioblastoma on chip

The BBTB associated with GBM has been recapitulated using a double-layered, compartmentalized microengineered chip model. The upper vascular channel was lined with a monolayer of human brain microvascular endothelial cells (HBMECs), while the lower tissue channel contained a 3D hydrogel-based network of human astrocytes (HAs) and patient-derived GBM cells (**Figure 1B** and **Supplementary Figure S1**). These two layers were separated by a porous membrane, mimicking the vascular-parenchymal interface. Continuous nutrient exchange and metabolic waste removal were maintained via side media channels integrated along the lower tissue channel (**Figure 1C**).

HBMECs cultured in the vascular channel formed a compact monolayer with continuous tight junctions (**Figure 1D**), while the co-cultured HAs and GBM cells established a 3D cellular network within 72 hours (**Figure 1E**). The BBTB model exhibited reduced barrier integrity, as indicated by increased permeability and reduced transendothelial electrical resistance (TEER), compared to the healthy BBB model (**Figure 1 F, G**). However, the presence of GBM cells did not significantly alter endothelial gene expression levels of junctional proteins, transporters, or receptors (**Supplementary Figure S3**), suggesting that our BBTB model recapitulate the structural preservation often observed at the infiltrative margins of GBM. These findings align with previous reports indicating that the endothelial barrier remains stable at the GBM margins compared to the barrier in the highly vascularized tumor core^[6,24,25]^.

### Crosstalk between glioblastoma and astrocytes

Astrocytes located in the peritumoral regions of GBM, commonly referred to as tumor-associated astrocytes, have been shown to promote the growth and progression of the tumor^[26,27]^, while GBM cells exploit these astrocytes to enhance their invasive potential and survival^[28]^. We co-cultured astrocytes with GBM cells isolated from GBM patient tissues to investigate these bi-directional interactions. Within the 3D microenvironment of our BBTB chip, both astrocytes and GBM cells exhibited branched and bushy shape, a physiological morphology observed in the brain-resident glial cells (**Figure 1E**). Notably, the GBM cells were uniformly and randomly distributed among astrocytes, rather than forming spheroids (**Figure 1E**). This distribution closely replicate the heterogeneous microenvironment observed at the GBM margins, where infiltrative GBM cells intermix with resident astrocytes^[29]^.

The interactions between astrocytes and GBM cells resulted in the significant upregulation of gene expressions of reactive gliosis markers, such as vimentin (*VIM*) and lipocalin 2 (*LCN2*) in astrocytes (**Figure 2A**). Moreover, plasminogen activator, urokinase (*PLAU*), which encodes urokinase-type plasminogen activator (uPA), was elevated in astrocytes in response to GBM contact (**Figure 2A**). uPA enhances the invasive capabilities of GBM cells via the uPA-plasmin cascade, which increases the activation of metalloproteinases (MMPs)^[28]^. In parallel, GBM cells co-cultured with astrocytes, showed the upregulated expression of transforming growth factor beta 1 (*TGF-β*), EPH Receptor A2 (*EphA2*), and vascular endothelial growth factor A (*VEGFA*) (**Figure 2B**), all of which contribute to invasive capacity of GBM^[30,31]^ and pro-angiogenic capability^[26]^. In particular, *EphA2* has been identified as a prognostic marker for GBM^[32,33]^. Taken together, these results demonstrate a dynamic crosstalk between GBM cells and astrocytes, in which tumor-derived signals activate astrocytes toward a reactive, tumor-promoting phenotype, while astrocytes reciprocally enhance the invasive and pro-angiogenic potential of GBM cells^[28,34]^.

**Figure 2.**
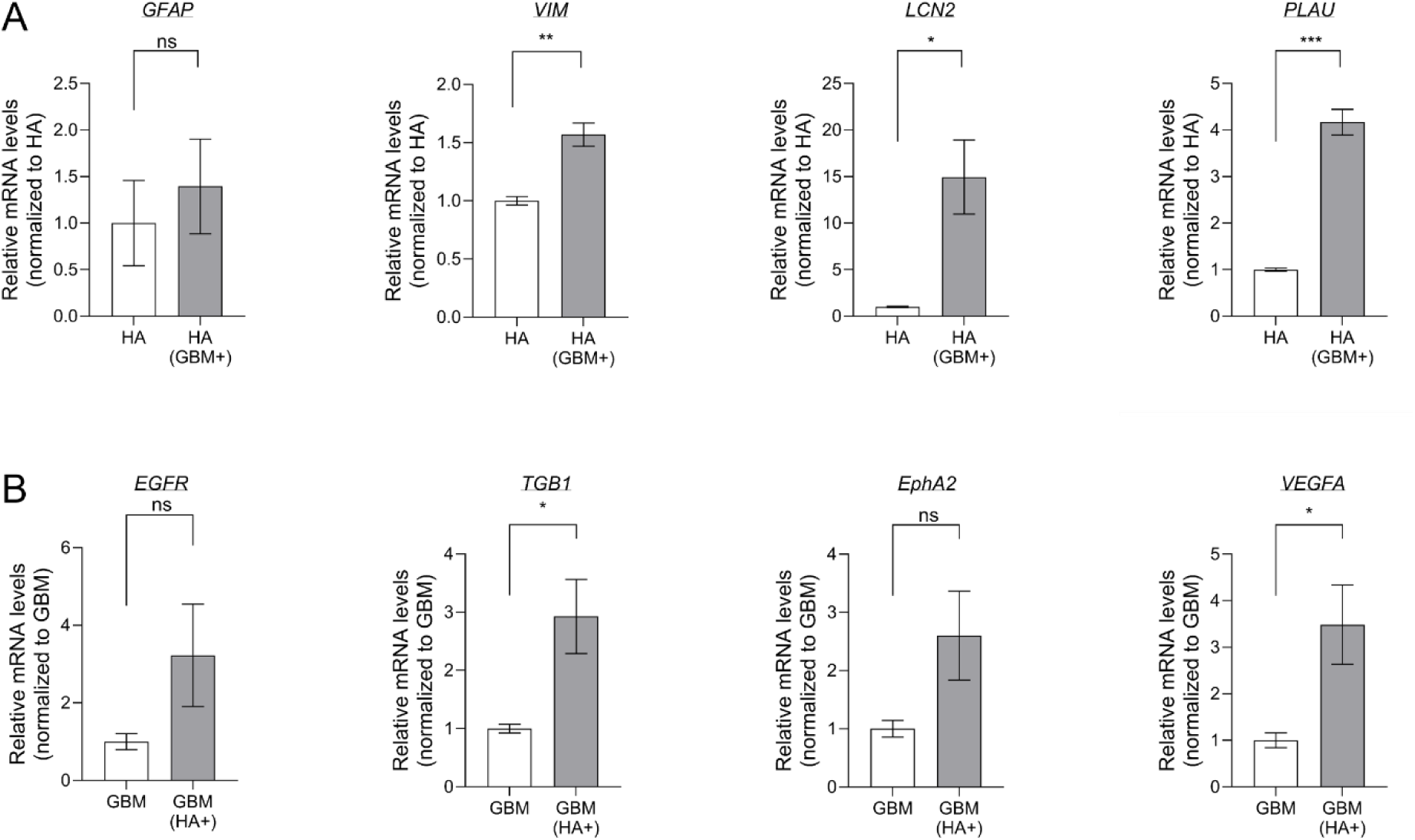
Interaction between HAs and GBMs. **(A)** Changes in gene expressions of HAs when co-cultured with GBM cells and **(B)** changes in gene expression of GBM cells when co-cultured with HAs, suggesting that HAs enhances tumor progression and invasiveness in the GBM environment. (n=3 for each condition, *p<0.05, **p<0.01 and ***p<0.001 by student’s t-test).

### Patient-specific blood-brain tumor barrier models

The BBTB represents the initial and critical obstacle to drug delivery in GBM, and its integrity profoundly influences the effectiveness of systemic chemotherapy. GBM has been shown to alter the integrity of the BBTB, however, the extent of this disruption is highly heterogeneous among patients. To investigate inter-patient variability of GBM, we established personalized BBTB models using GBM cells isolated from three patients with distinct molecular profiles (**Figure 3A, -C**). We first assessed barrier integrity using TEER and endothelial gene expression analysis. Among the three models, only patient A’s model (BBTB-A) exhibited a statistically significant reduction in TEER compared to the healthy BBB control, suggesting compromised barrier function. TEER values in models of patient B (BBTB-B) and C (BBTB-C) were comparable to the BBB model and not significantly different (**Figure 3D**). Consistently, gene expression analysis of endothelial junctional markers revealed that only BBTB-A downregulated tight junctional protein 1 (*TJP1*) and occludin (*OCLN*) relative to the BBTB-B and BBTB-C (**Figure 3E**). These results imply that the BBTB-A constructs a severely disrupted barrier, while the other two models (BBTB-B and BBTB-C) maintain relatively preserved vascular integrity. Notably, all models were constructed using the same endothelial cells (HBMECs) and platform, suggesting that the differences in barrier function arise solely from patient-specific GBM cells. This highlights the strong modulatory influence of tumor-derived factors on BBTB integrity.

**Figure 3.**
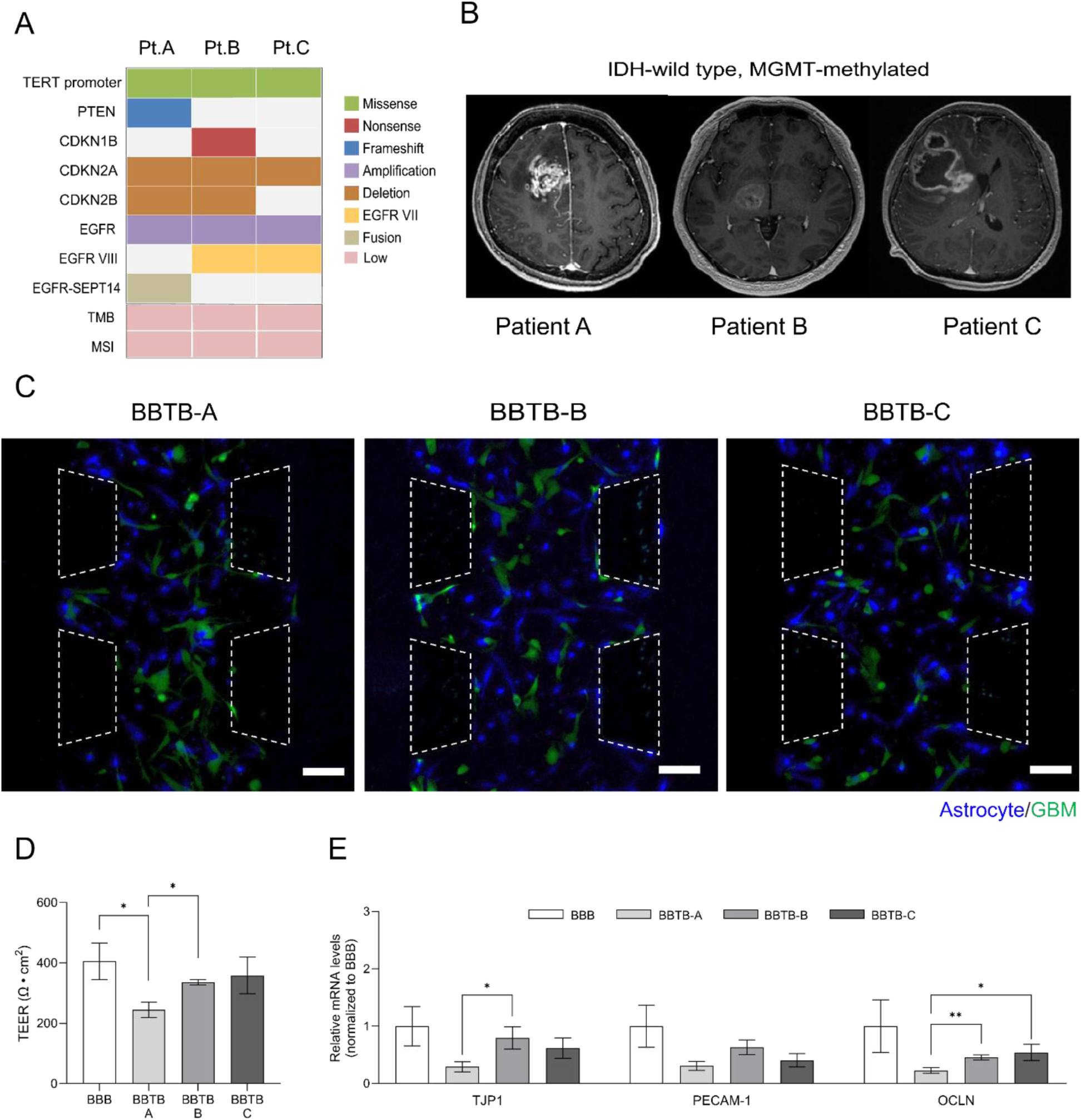
Establishment of the patient-specific BBTB model. **(A)** Oncoplot summarizing small variants, copy number variations (CNVs), gene fusions, tumor mutational burden (TMB) and microsatellite instability (MSI) identified through panel sequencing. (**B)** Representative magnetic resonance images of glioblastoma patients showing contrast-enhancing lesions. **(C)** Co-culture of astrocytes and GBM cells from different patients in our device (HAs, blue; patient-derived GBM cells, green) (lower channel, All scale bars = 100 µm). **(D)** TEER measurements comparing BBB and BBTB models (Patients A–C). **(E)** Relative mRNA expression of endothelial junction-related genes (*TJP1*, *PECAM-1*, *OCLN*) in patient-specific BBTB models, normalized to BBB controls (n=5 for BBB, BBTB-A, BBTB-B, and n=4 for BBTB-C, *p<0.05 and **p<0.01 by student’s t-test).

### Therapeutic responses in patient-specific BBTB models

All tumors were MGMT-methylated and IDH-wild-type tumors, indicating their potential sensitivity to alkylating chemotherapy agents like TMZ (**Figure 3B**). The efficacy of TMZ, the prime chemotherapy for GBM, has been known to be enhanced in MGMT promotor methylated tumors, by preventing tumor cells from repairing the drug-induced DNA damage^[35]^. However, the presence of diverse characteristics within GBM tumors can still lead to markedly different therapeutic outcomes even in patients with similar biomarker profiles. We confirmed that TMZ treatment induces tumor cell death on three distinct patient-specific models, while the responses varied among the models. Differential drug sensitivities were observed across the models (**Figure 4A**). In particular, BBTB-A and BBTB-B showed a significant increase in tumor cell death at a concentration of 50 µM TMZ, whereas BBTB-C did not show evident response. Moreover, BBTB-B and BBTB-C demonstrated sensitivity to TMZ between 50 µM and 500 µM, while BBTB-A showed negligible additional sensitivity (**Figure 4B**). These results indicate patient-specific variations in drug sensitivity across clinically relevant TMZ concentrations.

**Figure 4.**
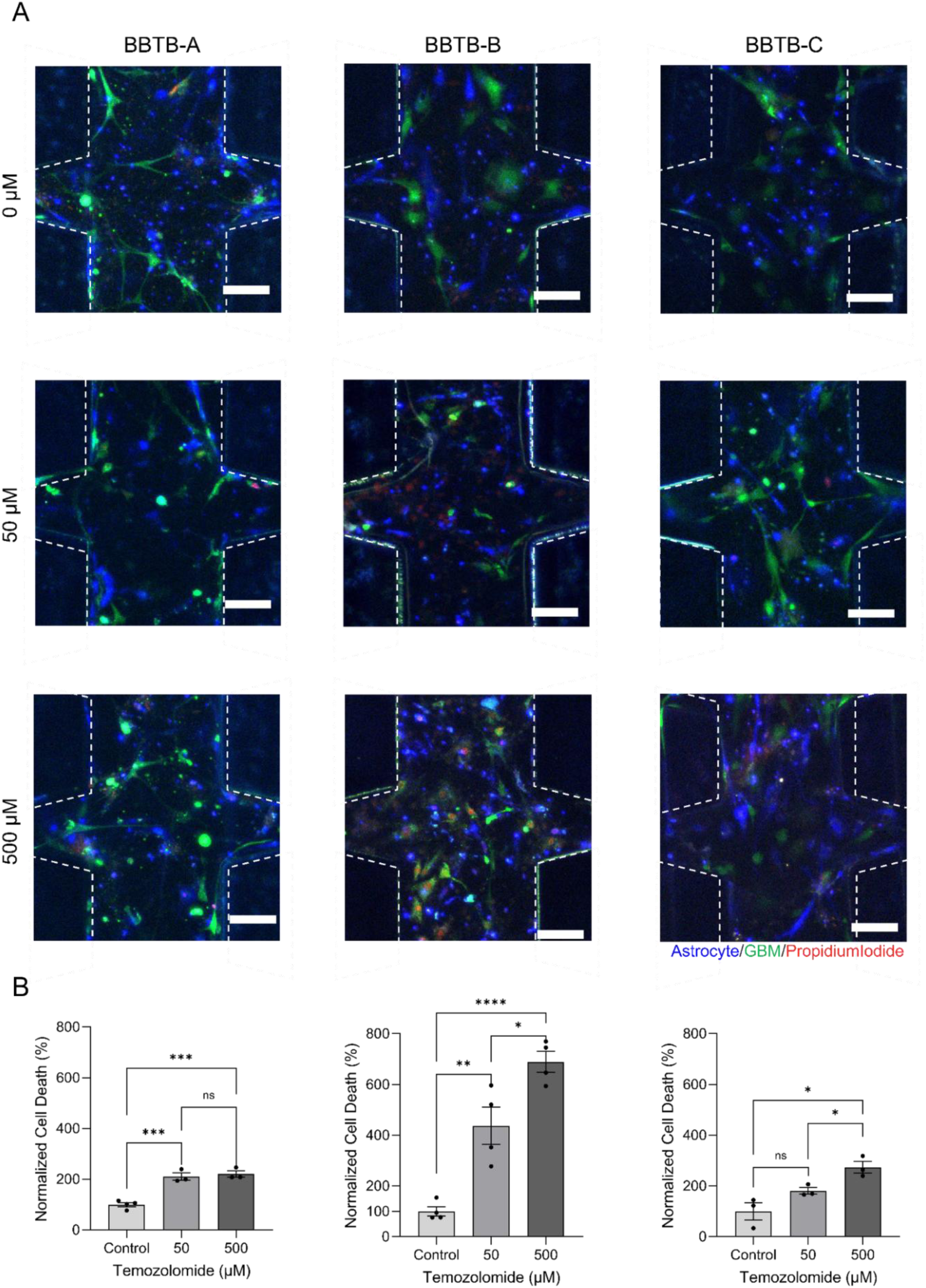
Chemotherapeutic responses in patient-specific BBTB models. **(A)** Dose-dependent viability of glioblastoma cells in BBTB chips from three different patients (Patients A–C) treated with 0, 50, or 500 µM TMZ. (HAs, blue; patient-derived GBM cells, green; PI (propidium iodide)-stained cells, red) (All scale bars = 100 µm). **(B)** Quantification of PI fluorescence intensity normalized to control for each patient-derived BBTB model under TMZ treatment (n ≥ 3 per group, *p<0.05, **p<0.01 and ***p<0.001 by student’s t-test).

Taken together, the patient-specific BBTB models revealed distinct relationships between vascular barrier function and chemotherapeutic response (**Figure 3D, 3E and 4B**). BBTB-A demonstrated high TMZ sensitivity at low concentrations, potentially due to its markedly disrupted vascular barrier, which may facilitate drug penetration. In contrast, BBTB-B exhibited strong drug responsiveness despite maintaining an intact barrier, suggesting inherent tumor sensitivity independent of vascular permeability. These findings underscore the importance of evaluating both tumor-intrinsic drug sensitivity and barrier integrity when assessing chemotherapeutic efficacy in GBM.

Notably, the results based on our BBTB models aligned well with clinical observations (**Figure 5A**). Patient A, whose model exhibited both a disrupted barrier and minimal TMZ response at high dose, exhibited the shortest progression-free survival (PFS) and overall survival (OS) (**Figure 5B**). Conversely, patient B, who showed strong drug sensitivity despite intact barrier function, showed the most favorable prognosis, including prolonged PFS and OS (**Figure 5B**). These correlations demonstrate the translational relevance of our personalized BBTB model and its potential to capture clinically meaningful patient-specific responses.

**Figure 5.**
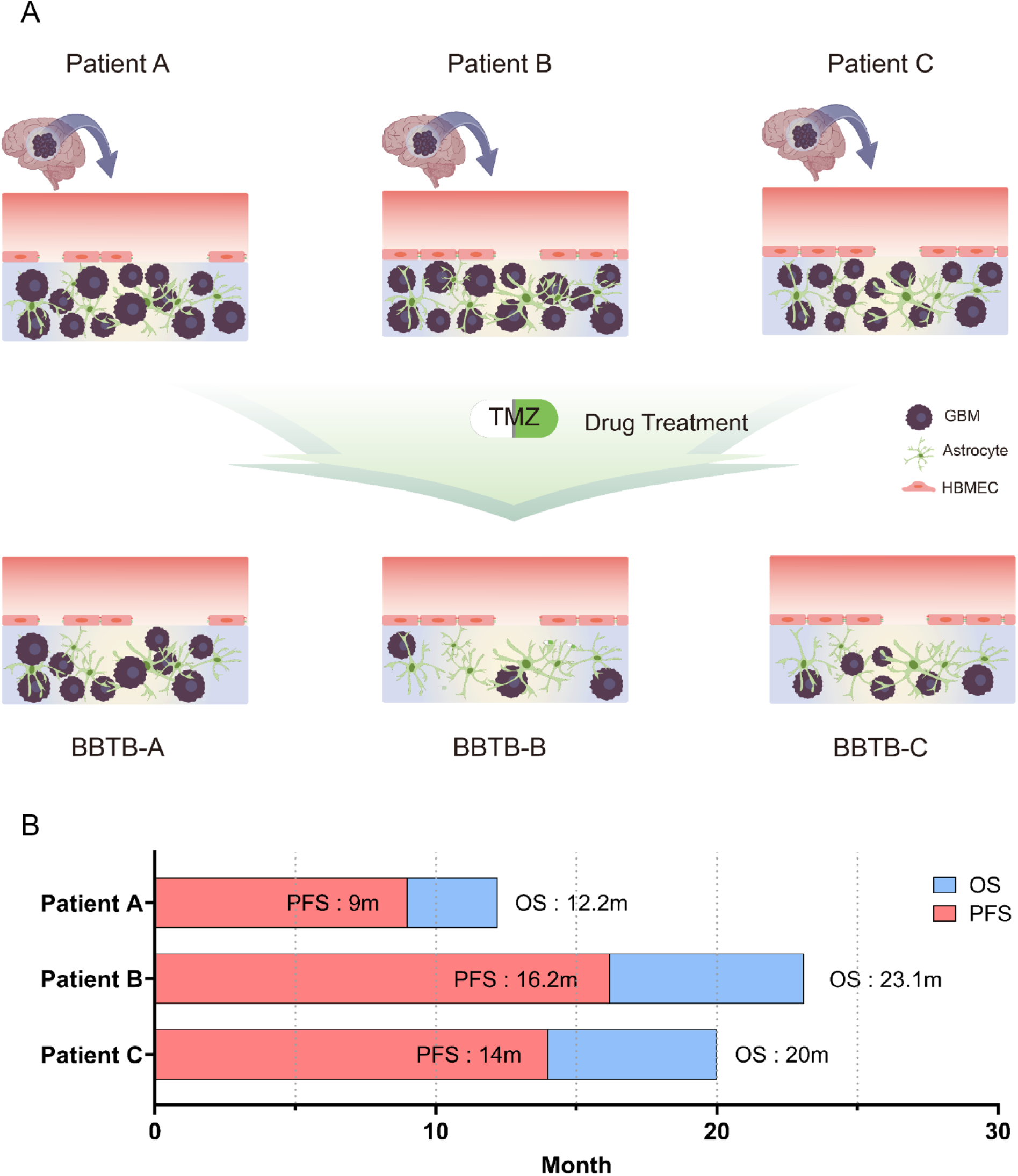
Overview of patient-specific variation in the BBTB model. **(A)** Agreement between on-chip findings and characteristics of the corresponding patient-derived GBM, suggesting the model’s ability to reflect individual tumor traits. **(B)** Swimmer plot illustrating progression-free survival (PFS) and overall survival (OS) for each patient. PFS (red) represents the duration before disease progression, while OS (blue) indicates the total survival period. Survival durations are shown in months (m).

## Discussion

In this study, we developed a microengineered BBTB-on-a-chip model that reconstructs the GBM margin, where the BBTB serves as the critical checkpoint for therapeutic agents^[36,37]^. As this barrier plays a decisive role in regulating drug penetration into infiltrative tumor regions, its physiological relevance is particularly important in the context of GBM treatment. To replicate this unique microenvironment, our platform integrates HBMECs, HAs, and patient-derived GBM cells within a double-layered design. This configuration facilitates the formation of a tight-junction-forming endothelial monolayer and establishes a 3D perivascular niche, thereby enabling direct interactions between tumor and stromal cells while preserving key features of the native BBTB.

The margin zone of GBM, which lies between healthy brain tissue and the GBM core, is extremely challenging for identification and surgical removal. Despite its lower density of tumor cells compared to the GBM core, the infiltrative characteristics of GBM cells contribute to recurrence and progression of GBM ^[38,39]^. Moreover, the GBM margin acts as a critical barrier for drug delivery in tumors due to its less disrupted BBTB ^[40]^ and a significant diffusion barrier ^[4]^. The interaction between normal astrocytes and GBM cells induces reactive gliosis, which is strongly associated with the diffusion properties of GBM and tumor progression ^[26,41]^. We have demonstrated increased reactive gliosis and invasive capacity of GBM cells through their interactions by examining the expression levels of the related genes.

In the GBM margin microenvironment, the integrity of BBTB is weakened, and such physical disruption can influence drug permeability. This addresses that drug efficacy is not solely determined by the intrinsic sensitivity of tumor cells, but also by the accessibility of therapeutic agents across the vascular barrier. BBTB disruption is neither uniform nor complete where certain regions may display leaky vasculature, others retaining intact tight junctions. In addition to this spatial heterogeneity, inter-patient heterogeneity further contributes to diverse barrier integrity, with some patients showing highly permeable BBTB and others exhibiting relatively preserved barriers. As drug permeability among patients^[42]^, patient-specific assessment of barrier integrity may enable personalized treatment planning. We evaluated the patient’s barrier integrity by measuring TEER across the patient BBTB models. TEER data confirmed substantial heterogeneity in barrier integrity among patients. Notably, one case exhibited a profoundly disrupted BBTB among three patients. This was further supported by the reduced expression of endothelial junctional markers, including *TJP1*, *PECAM-1*, and *OCLN*.

Together with assessing barrier integrity, we also established TMZ responsiveness in each patient’s BBTB model. Although all three patients were diagnosed with IDH-wildtype GBM and exhibited MGMT promoter methylation, our model revealed considerable differences in TMZ responsiveness among their respective BBTB models (**Supplementary Figure S4**).

These findings emphasize that genomic markers alone may be insufficient to predict therapeutic outcomes. Expanding the use of patient-specific models to evaluate responses to a broader range of therapeutic agents could provide actionable insights for personalized therapy.

Our platform delivers results within two weeks post-surgery, offering an opportunity to guide personalized treatment early in the clinical timeline. While standard clinical markers are often insufficient to predict therapeutic responses and patient prognosis, this functional model could add an additional layer of biological relevance that may enhance clinical decision-making. There is ample room for advancement in further characterizing the pathological features by incorporating analyses such as protein expression and cytokine profiling. Altogether, we believe our model offers a valuable platform for exploring the GBM margin and the BBTB. This approach may further support the development of personalized GBM therapeutic strategies.

## Ethics Statement

The use of patient tissues was approved by the Institutional Review Board (IRB No. CHAMC2021-01-024-001) at CHA Bundang Medical Center, Republic of Korea, with written informed consent obtained from all patients.

## Authorship

M.S.R., S.J.C., and S.I.A. wrote the manuscript. M.S.R., J.H.A., G.E.L., S.J.C., and J.W.J. performed experiments. M.S.R., J.H.A., G.E.L., J.W.K, and S.J.C. analyzed data. G.E.L., S.H.H., and M.S.R. performed RT-qPCR experiments. M.S.R., J.W.J., and S.J.C. performed immunocytochemistry. M.S.R., J.W.J., and S.J.C. fabricated microfluidic devices. Y.J.K., J.M.S., and J.M.P. performed glioblastoma cell isolation and culture. J.H.A., S.I.A., Y.J.K., and J.J.L. conceived, initiated, and supervised the overall project.

## Data Availability

All relevant data supporting the findings of this study are available within the paper and supplementary information files, and other data that support this study are available from the corresponding author upon reasonable request.

---

**Supplementary figure S1.**
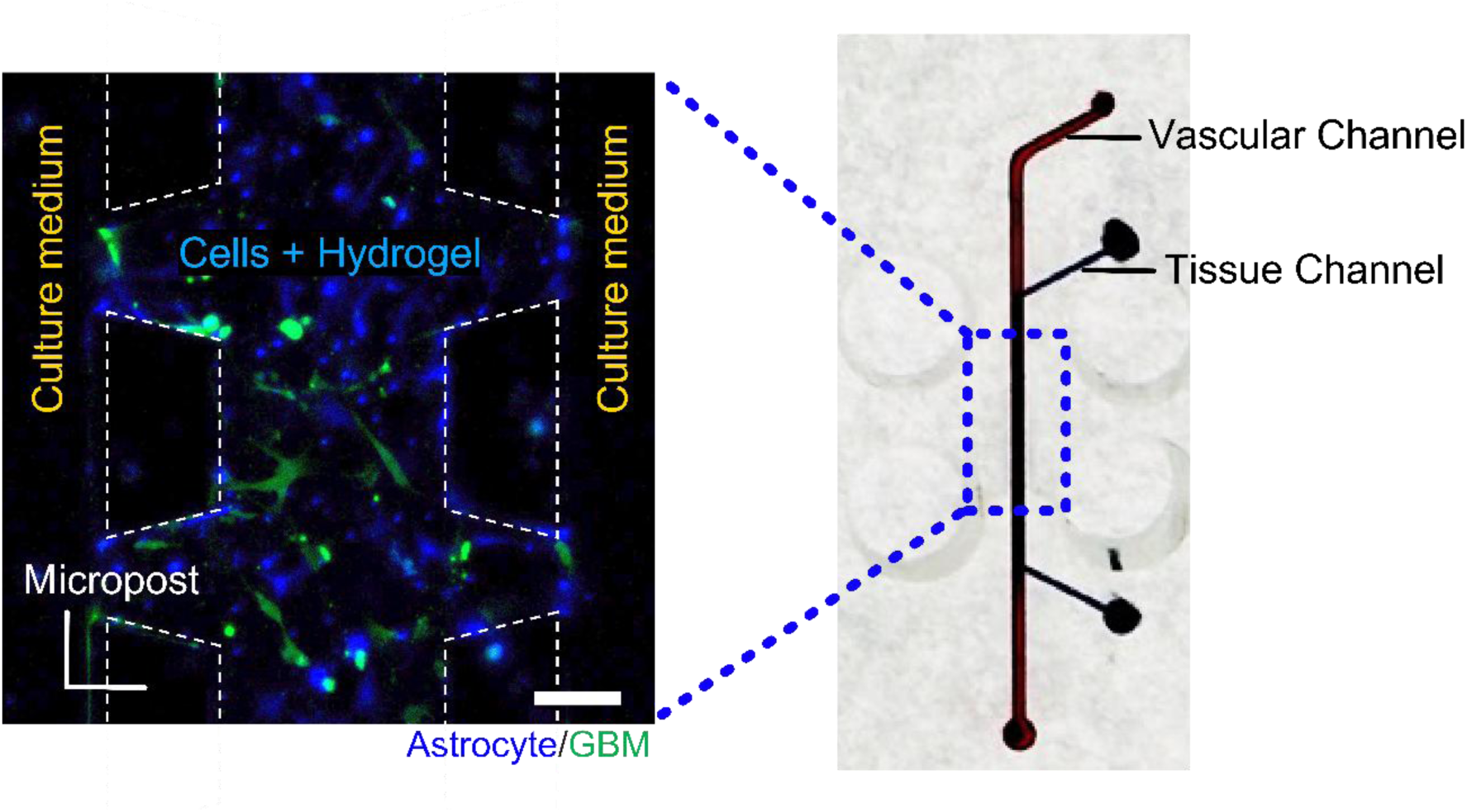
A photo of the fabricated device. The upper channel of the device made of PDMS is marked with red ink and the lower center channel is marked with blue ink (right). A magnified view of the lower channels (left) shows cells in hydrogel in the lower center channel bordered by microposts and culture medium in the lower side channel (scale bar = 100 µm).

**Supplementary figure S2.**
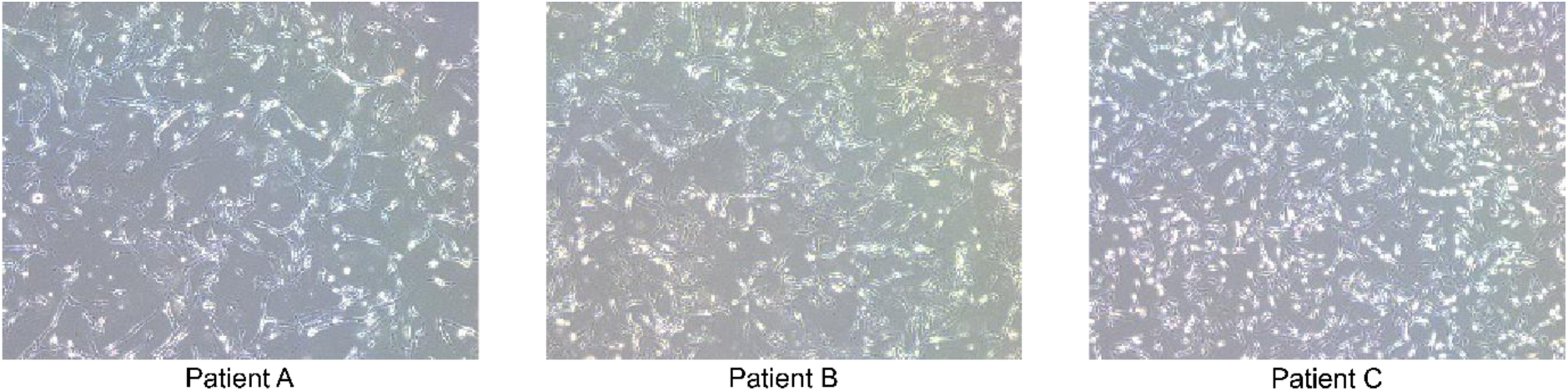
Bright field images of glioma cells from GBM patients utilized in this study.

**Supplementary figure S3.**
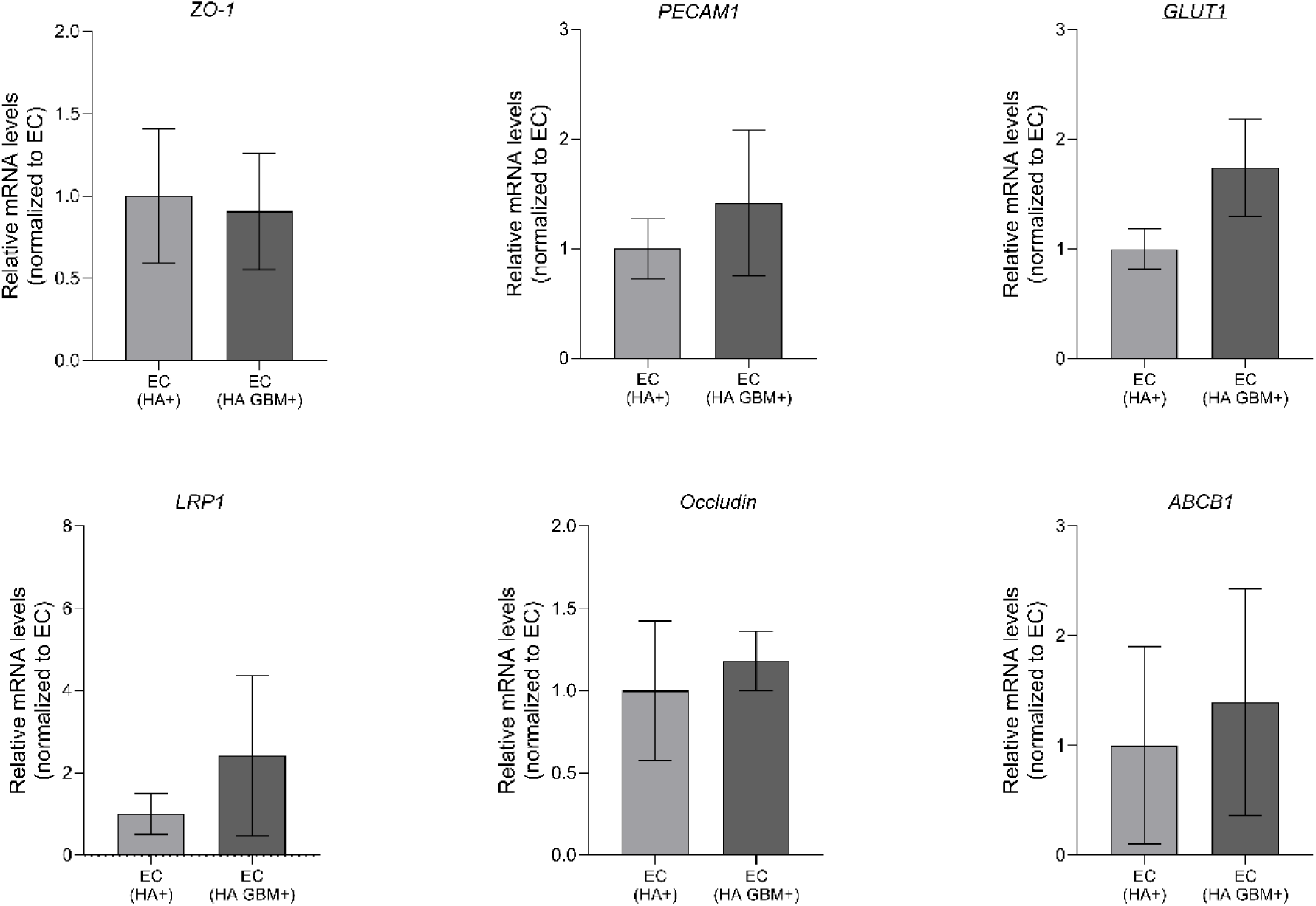
Comparison of relative mRNA levels in endothelial cells under mono- and co-culture conditions. (n=3 for each condition, *p<0.05, **p<0.01 and ***p<0.001 by student’s t-test).

**Supplementary figure S4.**
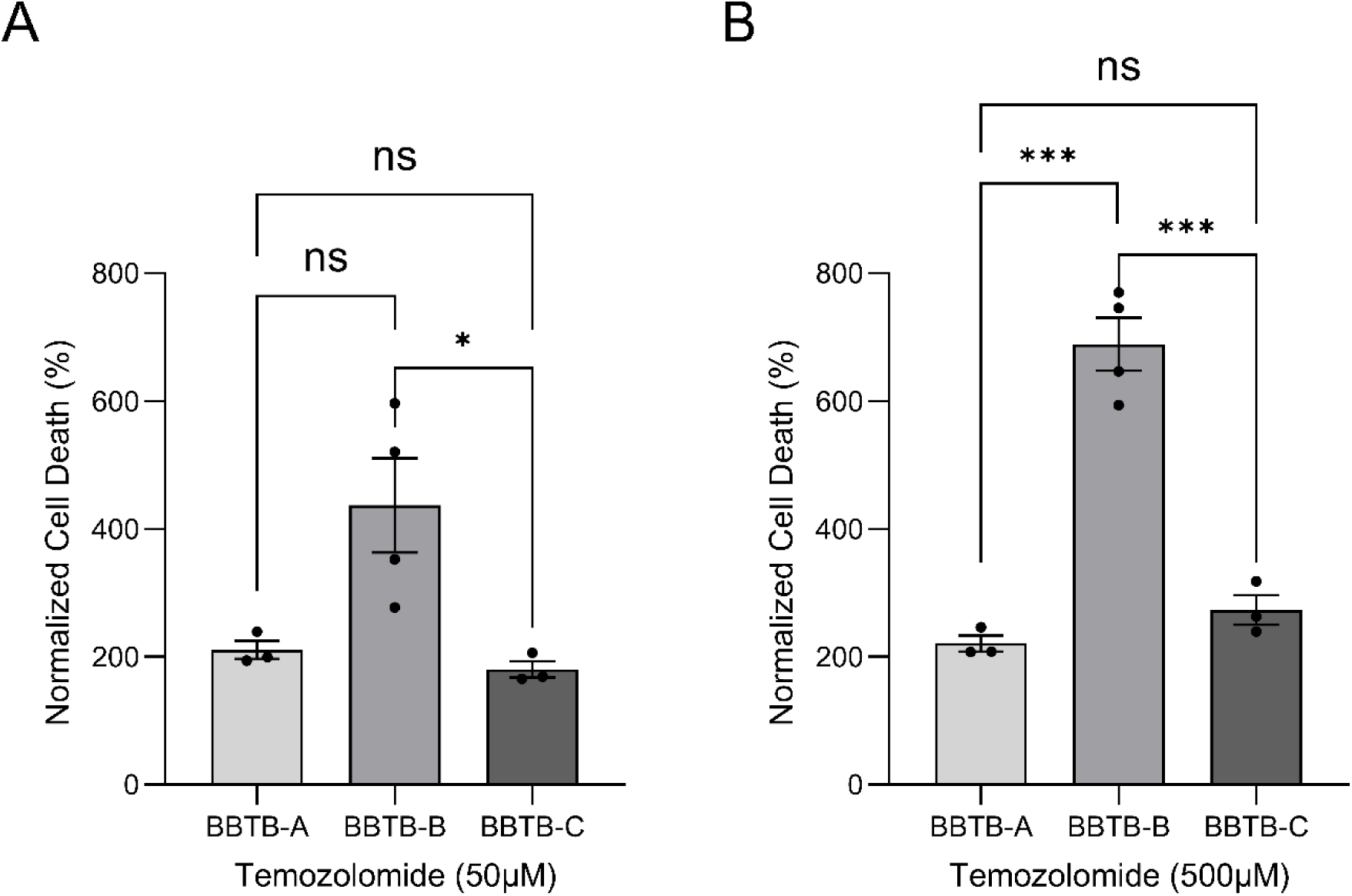
Normalized cell death rate of patient-derived GBM cells after TMZ treatment, relative to baseline response at 0µM.

**Supplementary Table S1.**
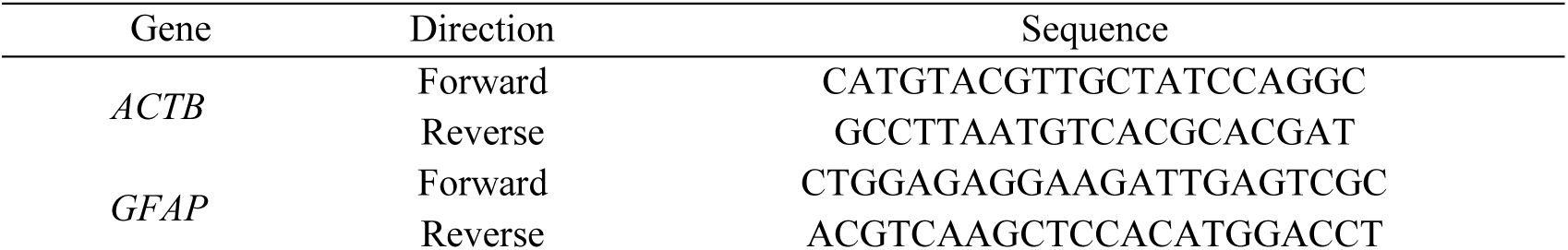
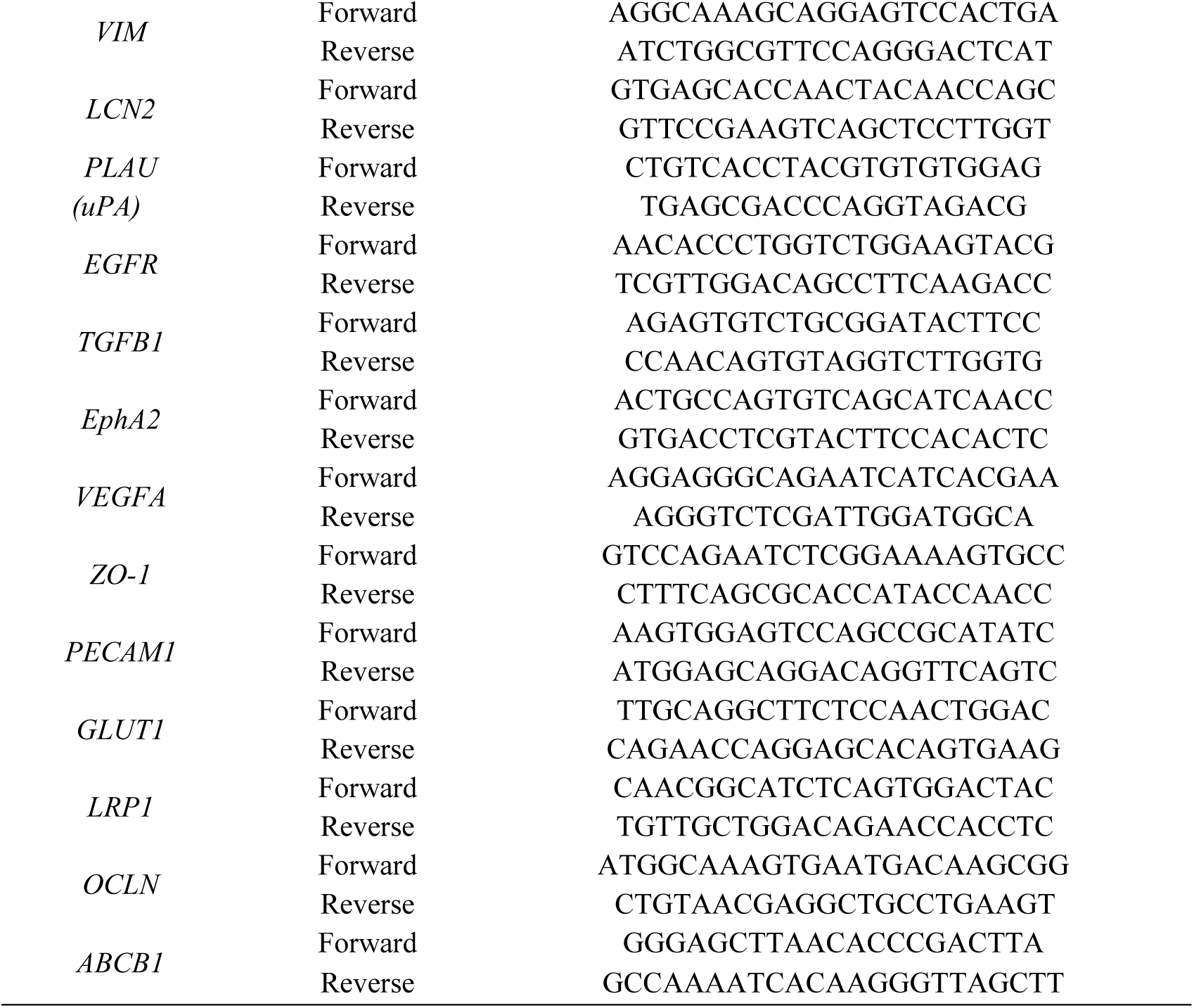
Primer sequence pairs used for qRT-PCR.

**Supplementary Table S2.**
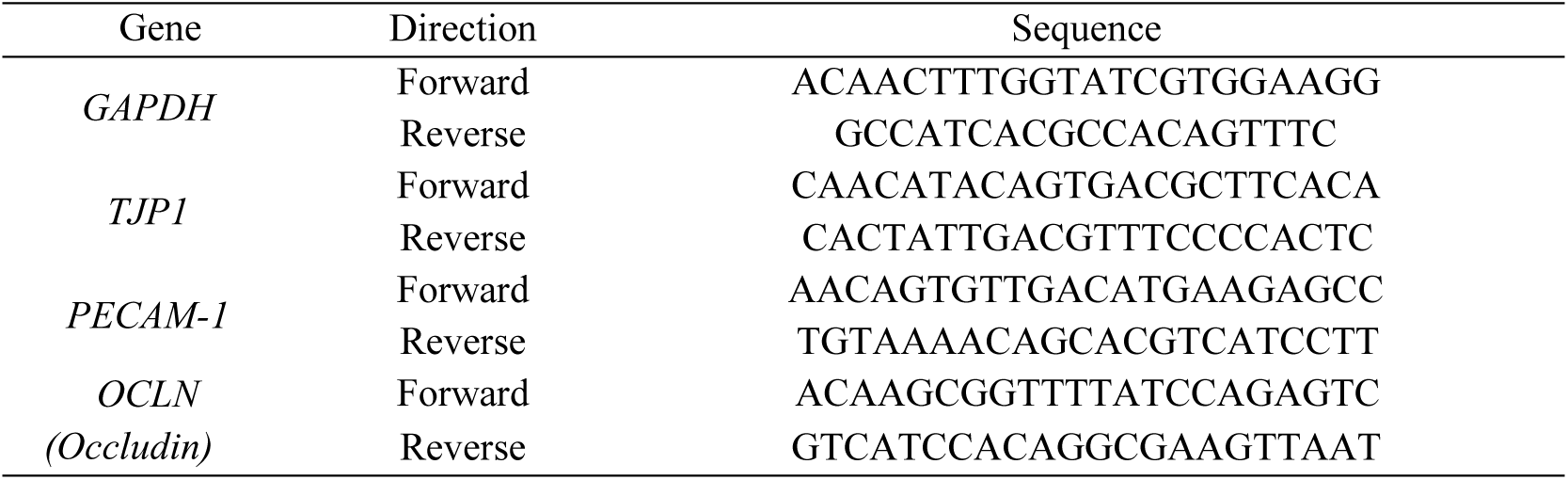
Primer sequence pairs used for patient specific qRT-PCR.

